# Lived Experiences Following Sepsis: Burden, Challenges, and Care Needs - A Qualitative Study

**DOI:** 10.64898/2026.09.16.26363207

**Authors:** Nandakumar Ravichandran, Diarmuid Quinlan, John Broughan, Ellen Hayes, Saswati Ghosh, Patricia Fitzpatrick, Nia Clendennen, Walter Cullen

## Abstract

**Background:** Sepsis is a life-threatening medical condition caused by the body’s extreme response to an infection. It can have lasting physical, cognitive, psychological and social consequences that extend beyond the acute illness. However, limited research has explored how people with a history of sepsis and their families experience these longer-term consequences and engage with healthcare services in Ireland.

**Methods:** A qualitative study was conducted using semi-structured interviews with 21 participants, including 14 people who had personally experienced sepsis and seven family members of people affected by sepsis. Participants were recruited through professional networks and a sepsis charity organisation in Ireland. Interviews were conducted by telephone or Zoom between May and June 2026, audio-recorded and transcribed verbatim. Data were analysed using Braun and Clarke’s reflexive thematic analysis.

**Results:** Five themes were identified: (1) recovery - a prolonged journey of physical & cognitive rebuilding; (2) navigating a recovery journey: lack co-ordination and integration; (3) continuity of support was uneven and often depended on individual healthcare professionals; (4) recovery reshaped family roles, work and everyday life; and (5) participants suggested a more coordinated approach to recovery. Participants described persistent physical, cognitive and psychological difficulties, alongside substantial effects on family responsibilities, employment and everyday activities. While some participants experienced valuable support from individual healthcare professionals, many described fragmented services, limited information about recovery and uncertainty about where to seek ongoing support.

**Conclusion:** Individuals with a history of sepsis and their families described substantial and ongoing consequences and variable experiences of longer-term care. The findings highlight the need to better recognise and respond to longer-term health needs following sepsis and to develop coordinated, accessible approaches to ongoing support in Ireland.

## Background

Post-sepsis syndrome (PSS) describes a range of persistent physical, cognitive, and psychological problems that may occur following sepsis (1–3). It affects around 50% of people surviving sepsis and may include fatigue, muscle weakness, chronic pain, cognitive difficulties, anxiety, depression, and post-traumatic stress symptoms (1–4). These symptoms can persist for years and may affect physical functioning, employment, relationships, and everyday activities, contributing to reduced quality of life and continued healthcare needs (1–4).

PSS may affect individuals’ ability to return to work and resume usual activities, while family members may experience ongoing emotional and practical impacts associated with supporting someone after sepsis. Individuals may also require continued healthcare and rehabilitation, placing additional demands on health services (5–10). While increasing attention has been given to sepsis survivorship and PSS internationally (11), limited evidence exists regarding how people living with persistent post-sepsis problems and their families experience recovery and navigate healthcare, particularly in Ireland (12). Understanding these experiences is important for identifying unmet needs and informing the development of appropriate post-sepsis support.

This qualitative study explored the experiences of people experiencing persistent post-sepsis symptoms and their family members in Republic of Ireland, with particular focus on the burden of persistent symptoms, experiences of healthcare and follow-up, and priorities for improving post-sepsis care.

## Methods

### Study setting, participants and sampling

This study forms part of a broader qualitative research project examining experiences of individuals with a history of sepsis and their family members in Ireland. The dataset is also being used in a related manuscript, “Navigating Sepsis: Experiences of Patients and Family Members in Ireland - A Qualitative Study”. Although the studies draw on the same participants and interviews, they address distinct research questions and have different analytical focuses (see S1 Appendix). This qualitative study included 21 participants: 14 people who had previously experienced sepsis and seven family members of people affected by sepsis. Participants were recruited in Ireland between 30 April and 25 May 2026 through professional networks associated with University College Dublin School of Medicine and through a non-profit sepsis charity organisation Irish Sepsis Foundation (13). Recruitment information invited individuals with relevant personal or family experience of sepsis to contact the research team. Interested participants were provided with an information leaflet describing the study. Written informed consent was obtained before each interview.

### Methodological approach and research paradigm

This study was informed by a constructivist paradigm and guided by the Social Ecological Model (SEM) (14, 15). The study was situated within a constructivist paradigm, recognising that participants’ accounts of illness and recovery are shaped by their individual circumstances, relationships and interactions with healthcare and wider society. This approach was appropriate for examining how people understood their experiences during and after sepsis, including how they interpreted persistent symptoms, healthcare encounters and recovery (14). The SEM provided a sensitising framework for considering influences operating at different levels, from individual experiences and interpersonal relationships to community, healthcare-system and wider policy contexts. Rather than being used to categorise participants’ accounts in advance, the framework informed interpretation of how experiences of recovery and care were situated within these different contexts (15). The study was conducted and reported in accordance with the Standards for Reporting Qualitative Research (SRQR) guidelines (16).

### Data collection

Semi-structured interviews were conducted by NR between 29 May and 19 June 2026, using telephone or Zoom according to participant preference. Interviews were audio-recorded and transcribed verbatim. Transcripts were pseudonymised before analysis to protect confidentiality. The interview guide focused on participants’ experiences following sepsis, including persistent symptoms and their impact, understanding of recovery, interactions with healthcare services, follow-up and support, and views on how post-sepsis care could be improved. The interview guide is provided in Supplementary Appendix 1.

### Data analysis

Interview data were analysed in NVivo 12 using Braun and Clarke’s reflexive thematic analysis (17). Analysis was iterative and involved repeated engagement with the transcripts, development of initial codes, examination of patterns across the dataset, construction and refinement of themes, and interpretation of the meanings underlying participants’ accounts. NR led the analysis, with JB contributing to secondary review and discussion of developing interpretations. Reflexivity was maintained throughout the research process. The researchers conducted five meetings to refine the themes.#

### Ethical considerations

Ethical approval for this study was granted by the University College Dublin Human Research Ethics Committee (LS-26-08-Ravichandran-Cullen).

## Results

A total of 21 participants took part in the study, including 14 people with a personal history of sepsis and seven family members of people who had experienced sepsis or died following sepsis. The family members were not relatives of the 14 participants with personal experience of sepsis. Participants ranged in age from 34 to 71 years (mean 49.8 years); 18 were women (85.7%) and three were men (14.3%). Participants were recruited from across Ireland, with urban and rural experiences represented.

Reflexive thematic analysis generated five interrelated themes describing how participants experienced recovery after sepsis: (1) recovery - a prolonged journey of physical & cognitive rebuilding; (2) navigating a recovery journey: lack co-ordination and integration; (3) continuity of support was uneven and often depended on individual healthcare professionals; (4) recovery reshaped family roles, work and everyday life; and (5) participants suggested a more coordinated approach to recovery. Across the themes, participants described both supportive experiences and significant gaps in care, with the ability to navigate recovery often dependent on personal resources, family support and access to responsive healthcare professionals.

### 1. Recovery - a prolonged journey of physical & cognitive rebuilding

Participants described recovery as extending beyond the resolution of sepsis, often taking several years. Rather than experiencing discharge as the beginning of a prolonged period of rebuilding physical capacity, cognitive function and independence. The extent of this disruption varied between participants, but recovery was consistently described as an ongoing process rather than a clearly defined endpoint.

For some, physical recovery involved relearning basic functions. One participant described, *“I had to learn to walk again” (P4, F)*, while another described having *“lost every iota of muscle”* and rebuilding strength *“one square millimetre at a time” (P10, F)*. Others described having to regulate activity carefully because exceeding their physical limits could result in prolonged worsening: *“You have to really gauge where your limit is, because if you go beyond your limit, you end up crashing” (P5, F)*.

The consequences were not limited to physical function. Participants also described cognitive changes that were difficult to recognise and, in some cases, felt insufficiently documented. One participant contrasted their previous and current cognitive abilities: *“I would have had a very, very good recall. Whereas now I struggle a little bit” (P5, F)*. Another expressed frustration that these difficulties were not reflected in their medical records, stating, *“At no point does it mention my cognitive difficulties” (P13, F)*.

Although the burden was substantial, participants also described adaptation and gradual adjustment. One participant reflected, *“I’ve kind of learned to accept the way it is now” (P21, F)*, while another, three and a half years after sepsis, continued to describe themselves as recovering: *“I’m 3*.*5 years post sepsis and believe I’m still recovering” (P4, F)*. Thus, participants described recovery as a prolonged and uneven process of rebuilding physical and cognitive function, with the duration and impact of ongoing difficulties often extending beyond their expectations of recovery.

### 2. Navigating a recovery journey: lack co-ordination and integration

Participants’ experiences of healthcare after sepsis were characterised by a contrast between high levels of healthcare contact and limited continuity between services. Several participants had numerous appointments with different specialists, yet this did not necessarily translate into coordinated care.

One participant described the complexity of managing multiple services: *“you’re dealing with so many departments, it’s not just the sepsis” (P1, M)*. Another recalled extensive specialist involvement, including vascular, renal and neurological follow-up, describing *“loads of follow-up”* across different services (P14, F). However, the presence of multiple appointments did not necessarily mean that participants felt someone was overseeing their holistic recovery. As one participant put it, *“They don’t really look at the bigger picture” (P7, F)*.

This distinction helps reconcile participants’ apparently contradictory accounts of “loads of follow-up” alongside reports of “no follow-up”. The issue was not necessarily an absence of healthcare contact, but a lack of coordination and ownership across that contact. One participant stated, *“Integration is poor” (P10, F)*, while another described the problem more directly: *“There is no follow-up. The coordination is missing, actually” (P13, F)*.

Participants consequently described having to navigate the system themselves. One participant expressed frustration at repeatedly having to obtain referrals: *“Now I have to get a referral all over again*… *I’m a patient, I’m on record*… *this is ridiculous” (P6, F)*. Family members similarly identified gaps between hospital and primary care. One reported that their GP had received no information that their father had experienced sepsis and suggested that *“some kind of liaison maybe between the hospital and the GP”* was needed (P3, F).

Overall, participants’ experiences suggested that frequent contact with healthcare services did not necessarily translate into coordinated care, with responsibility for connecting services often falling to individuals themselves.

### 3. Continuity of support was uneven and often depended on individual healthcare professionals

Participants described marked variation in the support they received during recovery. Some recalled being discharged with little information about what to expect or where to seek help. One participant recalled, *“When I was discharged from there. Nothing. No one followed you” (P1, M)*. Another described discharge as *“okay, see you now, good luck”* in the absence of rehabilitation advice (P5, F).

The absence of structured guidance left some participants uncertain about what recovery should look like. One participant explained, *“Nobody prepared me and I was very shocked because I thought, okay, I’m recovered” (P10, F)*. Another described seeking information independently and receiving little guidance about available services. This uncertainty was compounded when participants felt that healthcare professionals did not fully understand their ongoing difficulties. One participant stated, *“I don’t think my GPs understand*… *being told you’re going to be okay wasn’t enough to get me through it” (P7, F)*.

In contrast, participants described particularly positive experiences when individual professionals provided explanation, reassurance and realistic expectations. One participant valued a GP who explained that feeling unwell could be part of recovery: *“He was saying you’re going to feel terrible. But this is normal. That’s what I wanted to know” (P7, F)*. Another recalled a GP contacting them on the evening of discharge and explicitly acknowledging the severity of their illness: *“You were actually really, really sick. This is not going to be easy to get over” (P16, F)*.

Other community professionals could also provide meaningful support. One participant felt that *“the podiatrist showed me more care and attention than anybody else in the medical system” (P1, M)*. However, access to such support was inconsistent. A participant described a public health physiotherapy appointment being cancelled twice before one home visit eventually took place (P3, F).

Thus, participants described continuity of support as uneven, with positive experiences often linked to individual healthcare professionals who provided explanation, reassurance and recognition of ongoing needs, rather than to a consistent recovery pathway.

### 4. Recovery reshaped family roles, work and everyday life

The effects of recovery extended beyond health and healthcare, altering participants’ roles within their families and their ability to work and participate in everyday activities. Participants described family members taking on additional responsibilities and, in some cases, experiencing emotional consequences alongside the person who had been ill.

One participant stated simply, *“It had huge impacts on the family” (P10, F)*. Another described the emotional burden of witnessing a relative’s illness, reflecting, *“Should I have done more, should I have kicked and screamed?” (P11, F)*. Parents described how recovery could affect their ability to resume previous family responsibilities. One participant described returning home exhausted and feeling that they had been *“put back into normal life, caring for all the kids” (P19, F)*.

Employment and career were similarly affected. One participant reflected, *“It did change my career totally, and life totally that way too” (P14, F)*. At the same time, participants described the importance of supportive employers in enabling recovery. One noted that their employer had been *“amazing”* and allowed them to take extended time away from work (P3, F).

Participants also described changes in their sense of identity and physical capability. One participant reflected on seeing other people running and stated, *“I still get very annoyed when I see people running. I want to run too” (P21, F)*. This suggests that the consequences of sepsis were not experienced through comparison with participants’ previous abilities and expectations for their lives.

Overall, participants described recovery as requiring them to renegotiate family, work and everyday roles, with support from family and employers facilitating adjustment, while ongoing limitations could make returning to previous roles difficult.

### 5. Participants sought a more coordinated approach to recovery

Participants’ suggestions for improvement were closely connected to the difficulties described across the other themes. Rather than describing a single intervention, participants identified a need for clearer information, continuity between services, and a recognisable point of support through which they could navigate recovery.

Several participants wanted greater public and professional understanding of the longer-term consequences of sepsis. One participant described awareness as a personal priority: *“My mission is people should be aware of sepsis” (P1)*. Others emphasised the need for clearer guidance about recovery, including *“guidelines*… *when it comes to post-sepsis recovery” (P5, F)*.

Participants also wanted somewhere to turn when they were unsure where to obtain help. One suggested *“some post-follow-up clinic who tells us where to go” (P5, F)*, while another described the potential value of *“something centralised that you could go to, one department or one consultant or one team” (P17, F)*. Importantly, these accounts do not necessarily imply that participants wanted a single physical service located in one part of Ireland. Rather, they expressed a desire for a clear, accessible route into appropriate expertise and support, regardless of where they lived.

Participants also identified peer and community support as potentially valuable. One suggested that recognised support groups, comparable with established organisations supporting other chronic conditions, could provide an important source of information and connection.

At the same time, participants demonstrated considerable agency in managing their own recovery. One participant described focusing on areas they could control: *“Who do I have control of? I’ve control of my own fitness, my own nutrition, and who I ask in healthcare” (P10, F)*. While this illustrates resilience and self-management, it also highlights the extent to which participants felt responsible for navigating a complex recovery process themselves.

Overall, participants described a need for coordinated, person-centred approaches to post-sepsis care that would support recovery, improve access to information, and reduce the burden of self-managing complex health needs.

## Discussion

### Key findings

This study explored the experiences of individuals with a history of sepsis and family members living with the longer-term consequences of sepsis in Ireland. Participants described recovery as an extended and often difficult process, characterised by persistent health and functional challenges, fragmented healthcare experiences, and uncertainty about where to obtain appropriate support. Although some participants described valuable support from individual healthcare professionals, many experienced limited continuity and coordination of care. Participants identified the need for clearer information, coordinated follow-up and accessible support throughout recovery.

### Comparison with existing literature

The substantial and persistent burden described by participants is consistent with national and international literature documenting ongoing physical, cognitive, psychological and social consequences following sepsis (8, 12, 18, 19). Participants’ accounts therefore reinforce evidence that recovery extends well beyond the sepsis and can substantially affect every day daily functioning. Difficulties accessing appropriate follow-up and rehabilitation were also consistent with previous research describing unmet needs among individuals with a history of sepsis and limited continuity of care after the acute episode (20). The uncertainty reported by participants regarding recovery and available support highlights the importance of clearer post-sepsis follow-up and information (20).

Participants’ experiences of fragmented healthcare and having to navigate multiple services themselves are consistent with international evidence describing poor coordination across healthcare services for post-sepsis recovery (21). The need for integrated and person-centred approaches to post-sepsis care has increasingly been recognised within the international literature (11). The findings therefore add an Irish perspective to existing concerns regarding continuity and integration of post-sepsis care.

The effects described on family life and employment are also consistent with literature demonstrating that the consequences of sepsis can affect family members and wider daily life (22). This supports a broader approach to recovery that recognises the needs of both individuals and those supporting them. Finally, the importance participants placed on supportive community and primary care professionals is consistent with evidence highlighting the role of ongoing healthcare support in recovery after sepsis (23). However, participants’ accounts also indicate that access to such support was variable.

### Strengths and limitations

A key strength of this study is the inclusion of both individuals with a history of sepsis and family members, allowing different perspectives on the longer-term consequences of sepsis to be explored. Participants were recruited from across Ireland and included experiences from both urban and rural settings. Recruitment through professional networks and a sepsis charity may have resulted in selection bias, with individuals with particularly strong experiences more likely to participate. The sample included substantially more women than men, which may limit the diversity of perspectives captured, particularly those of men. Some participants were recalling experiences that occurred several years previously, introducing the possibility of recall bias. As with qualitative research, the findings are intended to provide depth and contextual understanding rather than statistical generalisability.

### Implications for practice and policy

The findings highlight a need for clearer information about expected recovery and persistent post-sepsis symptoms, alongside improved communication between patients, families and healthcare professionals. Community and primary care services may have an important role in recognising and assessing ongoing post-sepsis problems, providing support and coordinating referral to rehabilitation or specialist services. Participants also identified structured follow-up to support recovery. Further research should examine how persistent post-sepsis symptoms can be consistently recognised and assessed, and evaluate models of follow-up and rehabilitation, including dedicated post-sepsis services. Research should also explore the information and support needs of family members. Improving discharge information and communication may provide opportunity to better prepare patients and families for recovery and to support earlier recognition of ongoing problems. Developing post-sepsis care pathways will require appropriate resourcing of community-based care, including funding and education to support primary care professionals in recognising and managing persistent post-sepsis symptoms. Public education and better communication across primary and secondary care may also improve awareness of the longer-term consequences of sepsis and available sources of support. Care pathways should be designed to provide equitable access to post-sepsis services across Ireland.

## Conclusion

Individuals with a history of sepsis and their family members described substantial longer-term consequences of sepsis and variable support during recovery. Participants identified fragmented care, follow-up, information and coordination, alongside valuable support from individual healthcare professionals. Developing clearer and more coordinated approaches to longer-term recovery may help address the unmet needs identified in this study.

## Acknowledgements

We sincerely appreciate the support provided by University College Dublin (UCD) School of Medicine including its Clinical Research Centre, College of Health and Agricultural Sciences and the UCD/HSE Dublin and South East GP Research Network.

## Supporting information

S1 Appendix. Topic guide

## Data reporting

The qualitative interview data generated and analysed during this study are not publicly available due to privacy and ethical restrictions, as the data contain detailed personal accounts of participants’ experiences of sepsis and healthcare. Given the small sample and the potentially identifiable nature of qualitative interview data, sharing the underlying transcripts could compromise participant confidentiality. Access to the underlying data may be considered on reasonable request to the corresponding author, subject to the relevant ethical, privacy and consent requirements.

## Notes

### Competing Interest Statement

The authors have declared no competing interest.

## References

1. Sepsis Alliance. Post-Sepsis Syndrome. 2026. Available from: https://www.sepsis.org/sepsis-basics/post-sepsis-syndrome/

2. The UK Sepsis Trust. Sepsis Recovery & Post Sepsis Syndrome. Available from: https://sepsistrust.org/get-support/sepsis-recovery-post-sepsis-syndrome/

3. Centers for Disease Control and Prevention. Managing Recovery from Sepsis. 2025. Available at: https://www.cdc.gov/sepsis/living-with/index.html

4. Chechulina V, Sheikh F, Lóser M, Englesakis M, Barrett K. Healthcare costs after sepsis: a systematic review. Critical Care. 2025;29(1):381.

5. Iwashyna TJ, Cooke CR, Wunsch H, Kahn JM. Population burden of long-term survivorship after severe sepsis in older Americans. Journal of the American Geriatrics Society. 2012;60(6):1070–1077.

6. Wang HE, Szychowski JM, Griffin R, Safford MM, Shapiro NI, Howard G. Long-term mortality after community-acquired sepsis: a longitudinal population-based cohort study. BMJ Open. 2014;4(1):e004283.

7. European Sepsis Alliance. Life After Sepsis Guide. 2020. Available at: https://www.europeansepsisalliance.org/guide#

8. van der Slikke EC, Beumeler LFE, Holmqvist M, Linder A, Mankowski RT, Bouma HR. Understanding Post-Sepsis Syndrome: How Can Clinicians Help? Infection and Drug Resistance. 2023;16:6493–6511.

9. Winters BD, Eberlein M, Leung J, Needham DM, Pronovost PJ, Sevransky JE. Long-term mortality and quality of life in sepsis: a systematic review. Critical Care Medicine. 2010;38(5):1276–1283.

10. Rahmel T, Schmitz S, Nowak H, Schepanek K, Bergmann L, Halberstadt P, et al. Long-term mortality and outcome in hospital survivors of septic shock, sepsis, and severe infections: The importance of aftercare. PLoS ONE. 2020;15(2):e0228952.

11. Draeger L, Fleischmann-Struzek C, Gehrke-Beck S, Heintze C, Thomas-Rueddel DO, Schmidt K. Barriers and facilitators to optimal sepsis care - a systematized review of healthcare professionals’ perspectives. BMC Health Services Research. 2025;25(1):591.

12. Fleming A, Barbosa TM, Smiddy M, O’Driscoll M, Murphy K, Murphy S, Heinrich C. A Patient and Staff Stories’ Project regarding Antimicrobial Resistance, Infection Prevention and Control, and Sepsis, knowledge, practices, experiences and perceptions. 2025.

13. Irish Sepsis Foundation. Irish Sepsis Foundation. Available from: https://www.sepsisfoundation.ie/

14. Chun Tie Y, Birks M, Francis K. Grounded theory research: A design framework for novice researchers. SAGE Open Medicine. 2019;7:2050312118822927.

15. Podgorski CA, Anderson SD, Parmar J. A biopsychosocial-ecological framework for family-framed dementia care. Frontiers in Psychiatry. 2021;12:744806.

16. O’Brien BC, Harris IB, Beckman TJ, Reed DA, Cook DA. Standards for reporting qualitative research: a synthesis of recommendations. Academic Medicine. 2014;89(9):1245–1251.

17. Byrne D. A worked example of Braun and Clarke’s approach to reflexive thematic analysis. Quality & Quantity. 2022;56(3):1391–1412.

18. Lei S, Ruan H, Zhao H, Feng Z, Wang J, Zhao G, et al. Long-term prognosis of sepsis survivors after hospital discharge: A systematic review and meta-analysis of observational studies. Journal of Infection and Public Health. 2026;19(5):103211.

19. Schade Skov C, Østervang C, Brabrand M, Lassen AT, Nielsen DS. How do sepsis survivors experience life after sepsis? A Danish qualitative study exploring factors of importance. BMJ Open. 2024;14(2):e081558.

20. Vahl F, Ullmann S, Draeger L, Kannengießer L, Pletz MW, Matthaeus-Kraemer CT, Fleischmann-Struzek C. “Lost in Transition”: Informational Needs of Sepsis Survivors and Their Relatives Across the Care Trajectory-A Qualitative Study. Journal of Clinical Medicine. 2025;15(1).

21. Apitzsch S, Larsson L, Larsson AK, Linder A. The physical and mental impact of surviving sepsis - a qualitative study of experiences and perceptions among a Swedish sample. Archives of Public Health. 2021;79(1):66.

22. Gallop K, Kerr C, Nixon A, Verdian L, Barney J, Beale R. A Qualitative Investigation of Patients’ and Caregivers’ Experiences of Severe Sepsis. Critical Care Medicine. 2014;43.

23. Taylor R, Vollam S, McKechnie SR, Shah A. Improving Outcomes in Survivors of Sepsis—The Transition from Secondary to Primary Care, and the Role of Primary Care: A Narrative Review. Journal of Clinical Medicine. 2025;14(8):2582.

